# Associations between dietary patterns and gene expression pattern in peripheral blood mononuclear cells: a cross-sectional study

**DOI:** 10.1101/2020.01.25.20018465

**Authors:** Jacob J. Christensen, Stine M. Ulven, Magne Thoresen, Kenneth Westerman, Kirsten B. Holven, Lene F. Andersen

## Abstract

**Background:** Diet may alter gene expression in immune cells involved in cardio-metabolic disease susceptibility. However, we still lack a robust understanding of the association between diet and immune cell-related gene expression in humans.

**Objective:** Our objective was to examine the associations between dietary patterns (DPs) and gene expression profiles in peripheral blood mononuclear cells (PBMCs) in a population of healthy, Norwegian adults.

**Methods:** We used factor analysis to define *a posteriori* DPs from food frequency questionnaire-based dietary assessment data. In addition, we derived interpretable features from microarray-based gene expression data (13 967 transcripts) using two algorithms: CIBERSORT for estimation of cell subtype proportions, and weighted gene co-expression network analysis (WGCNA) for cluster discovery. Finally, we associated DPs with either CIBERSORT-predicted PBMC leukocyte distribution or WGCNA gene clusters using linear regression models. All analyses were gender-stratified (n = 130 women and 105 men).

**Results:** We detected three DPs that broadly reflected *Western, Vegetarian*, and *Low carbohydrate* diets. CIBERSORT-predicted percentage of monocytes associated strongly and negatively with the *Vegetarian* DP in both women and men. For women, the *Vegetarian* DP associated most strongly with a large gene cluster consisting of 600 genes mainly involved in regulation of DNA transcription. For men, the *Western* DP inversely associated most strongly with a smaller cluster of 36 genes mainly involved in regulation of metabolic and inflammatory processes. In subsequent protein-protein interaction network analysis, the most important *driver genes* within these WGCNA gene clusters seemed to physically interact in biological networks.

**Conclusions:** DPs may affect percentage monocytes and regulation of key biological processes within the PBMC pool. Although the present findings are exploratory, our analysis pipeline serves a useful framework for studying the association between diet and gene expression.

## Introduction

Cardio-metabolic diseases are the main causes of death worldwide (1). They are mainly caused by life-long exposure to classical risk factors such as obesity, hypertension, dyslipidemia and dysglycemia (2). Diet affects these risk factors and thereby contributes to the rate of disease progression (3). Diet can also influence gene expression in immune cells directly, and so potentially affect cardio-metabolic disease susceptibility (4–6). However, we still lack a thorough understanding of the association between diet and immune cell-related gene expression.

Free-living humans consume a variety of foods in combination. To capture this variation meaningfully, we often define so-called dietary patterns (DPs). *A posteriori* DPs are data-driven; they are defined based on the co-consumption of foods in the population under study (7). Naturally, a posteriori DPs reflect local food culture and have high internal validity. As such, a posteriori DPs may constitute robust measures of global diet exposure, and could be used to strengthen the reliability of associations between diet and biomarkers within a population. This may be especially relevant in order to examine high-variance biomarkers, such as gene expression profiles.

Peripheral blood mononuclear cells (PBMCs) are directly involved in the underlying pathophysiology of cardio-metabolic disease (8). They represent a mixture of cells that are transiently part of a specialized niche in the circulation, of which some move to sites of inflammation. Affected by a number of input signals, PBMCs adapt to their environment; dietary metabolites, interleukins and chemokines, classical risk factors, and a host of other factors all influence the transcriptome of the PBMCs (9).

Many previous studies in humans that have associated diet with PBMC gene expression have used a classical gene expression-wide association (gxWA) strategy (10,11). The underlying correlation structure of the transcriptome, however, provides an opportunity to improve upon gxWA methods. Biologically-relevant dimensionality reduction algorithms, such as CIBERSORT and weighted gene co-expression network analysis (WGCNA), simplify whole-genome gene expression matrices into interpretable features (12,13). These methods also increase the signal-to-noise ratio and thereby robustness of the features, while they simultaneously reduce the multiple testing burden (14).

The objective of the present study was to examine the associations between a posteriori-defined DPs and derived gene expression features in PBMCs in a population of healthy, Norwegian adults. We hypothesized that DPs would associate with PBMC gene expression, and that the associations would point to specific biological mechanisms that potentially mediate the effects of diet on cardio-metabolic diseases.

## Subjects and methods

### Study design and participants

The present study is based on cross-sectional data from the screening visit of a randomized controlled dietary intervention, presented in detail elsewhere (15). We included all participants from whom we had both dietary assessment data and PBMC gene expression data, in addition to standard clinical and biochemical measurements. After excluding four participants with self-reported energy intake above 25 MJ/d, we included 235 participants in the analyses (n = 130 women, n = 105 men).

The subject characteristics are presented in **Table 1**. Briefly, the men were younger than the women, but had a more unhealthy body composition and subsequent clinical sequelae. Both genders had moderate hypercholesterolemia.

**Table 1.**
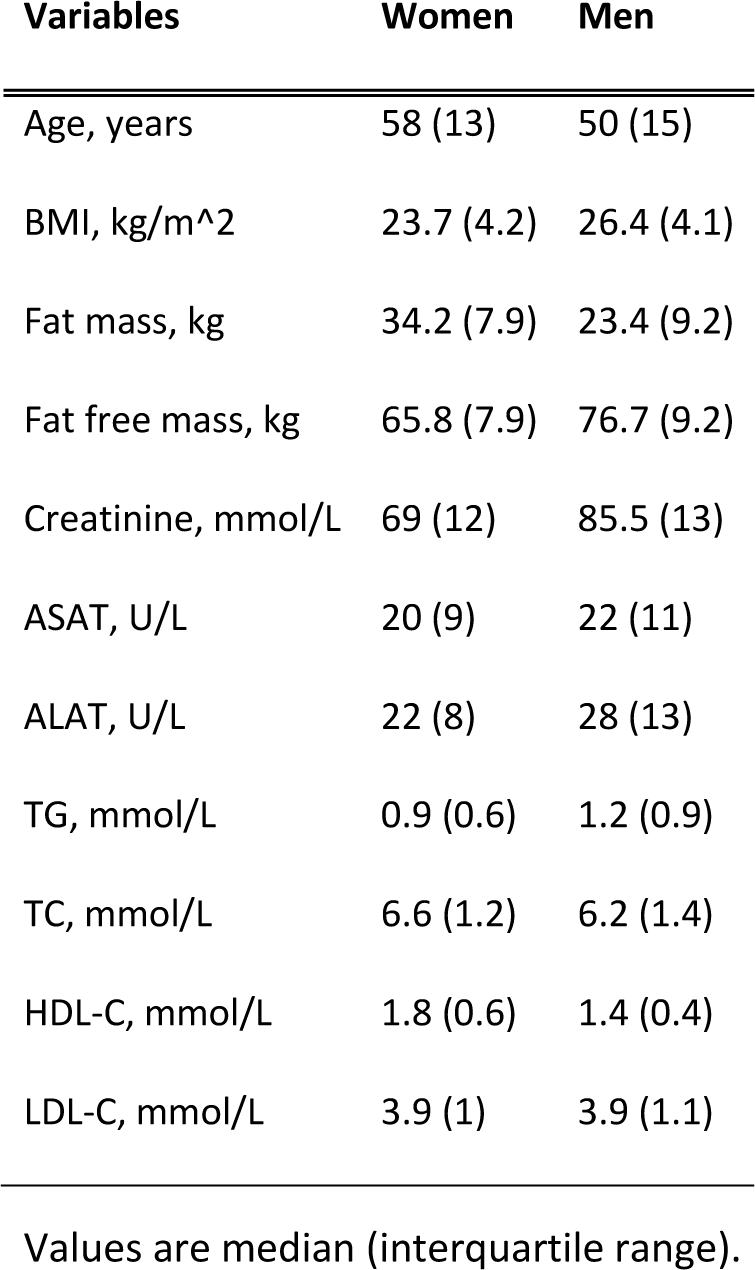
Study sample: clinical characteristics.

| Variables | Women | Men |
| --- | --- | --- |
| Age, years | 58 (13) | 50 (15) |
| BMI, kg/m<sup>2</sup> | 23.7 (4.2) | 26.4 (4.1) |
| Fat mass, kg | 34.2 (7.9) | 23.4 (9.2) |
| Fat free mass, kg | 65.8 (7.9) | 76.7 (9.2) |
| Creatinine, mmol/L | 69 (12) | 85.5 (13) |
| ASAT, U/L | 20 (9) | 22 (11) |
| ALAT, U/L | 22 (8) | 28 (13) |
| TG, mmol/L | 0.9 (0.6) | 1.2 (0.9) |
| TC, mmol/L | 6.6 (1.2) | 6.2 (1.4) |
| HDL-C, mmol/L | 1.8 (0.6) | 1.4 (0.4) |
| LDL-C, mmol/L | 3.9 (1) | 3.9 (1.1) |

Values are median (interquartile range).

### Data types

We used a food-frequency questionnaire (FFQ) to assess habitual food intake from the preceding year (16). From the originally 323 food items, we removed 41 items due to unclear interpretation, and grouped the remaining 282 into 33 food groups, based on food category and nutrient content (**Table 2**). Self-reported intake of foods and nutrients are presented in **Supplementary Table 1, Supplementary Table 2** and **Supplementary Table 3**. Furthermore, we collected PBMCs and extracted RNA according to standardized protocols, as previously described (6).

**Table 2.**
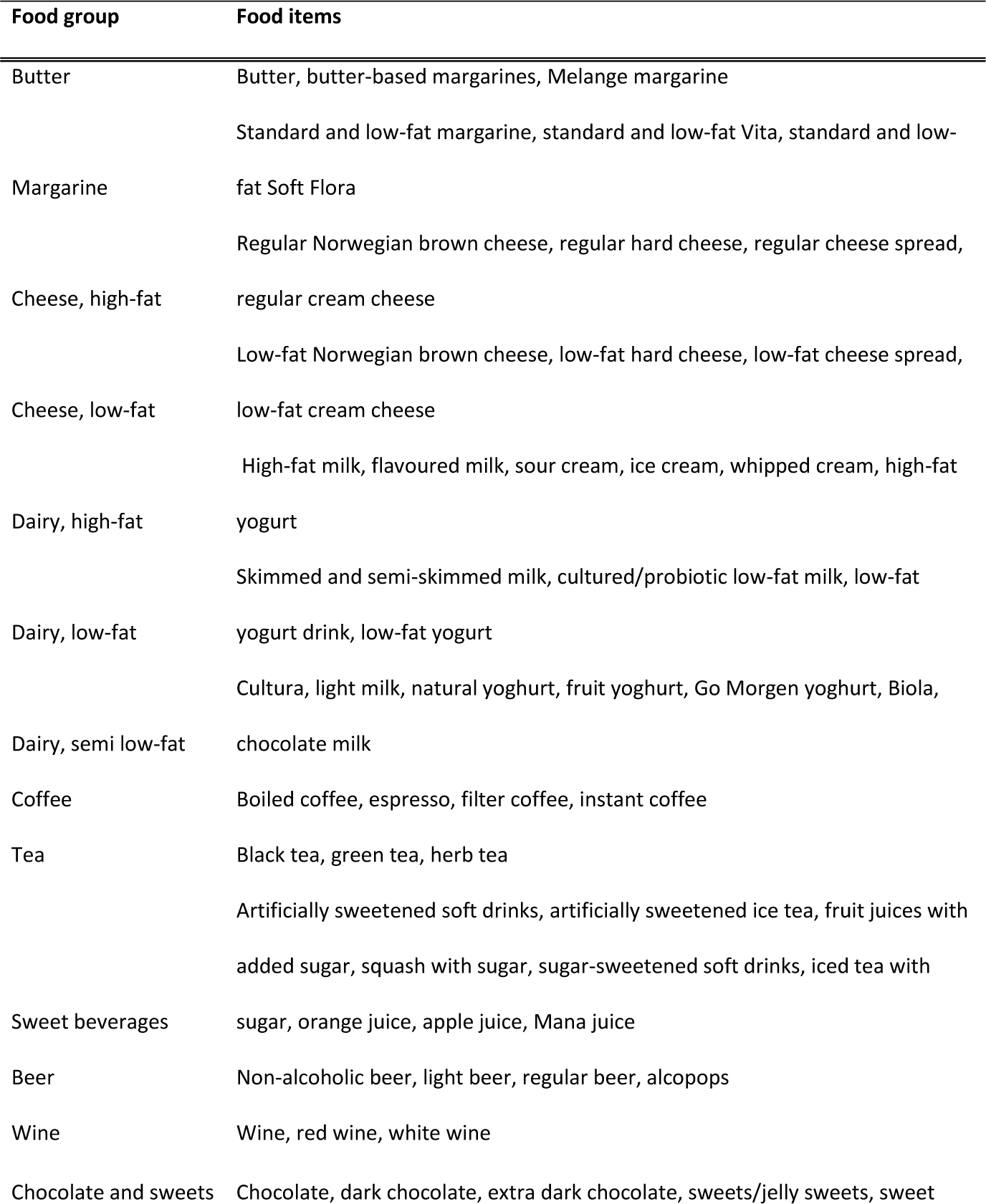
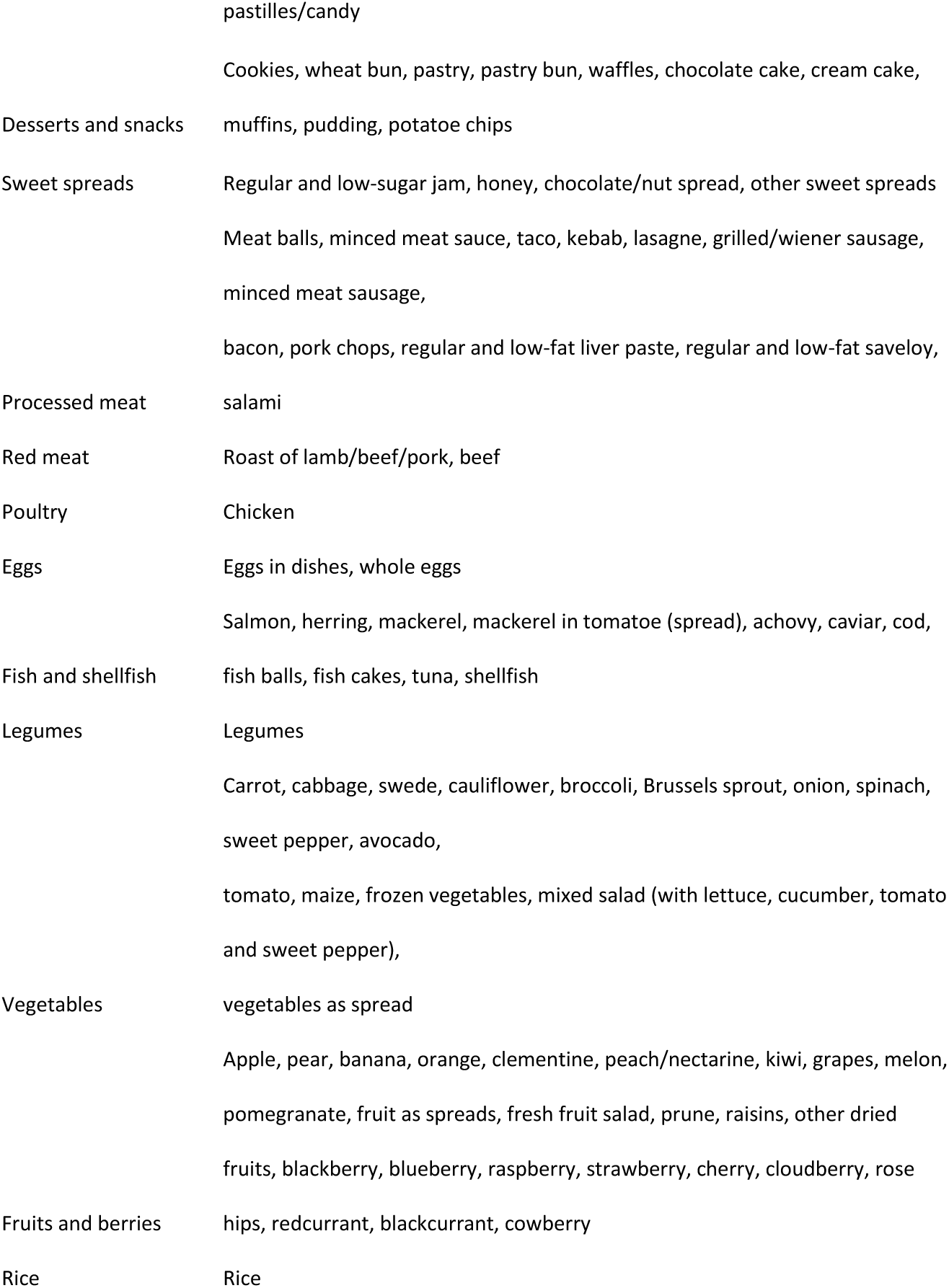
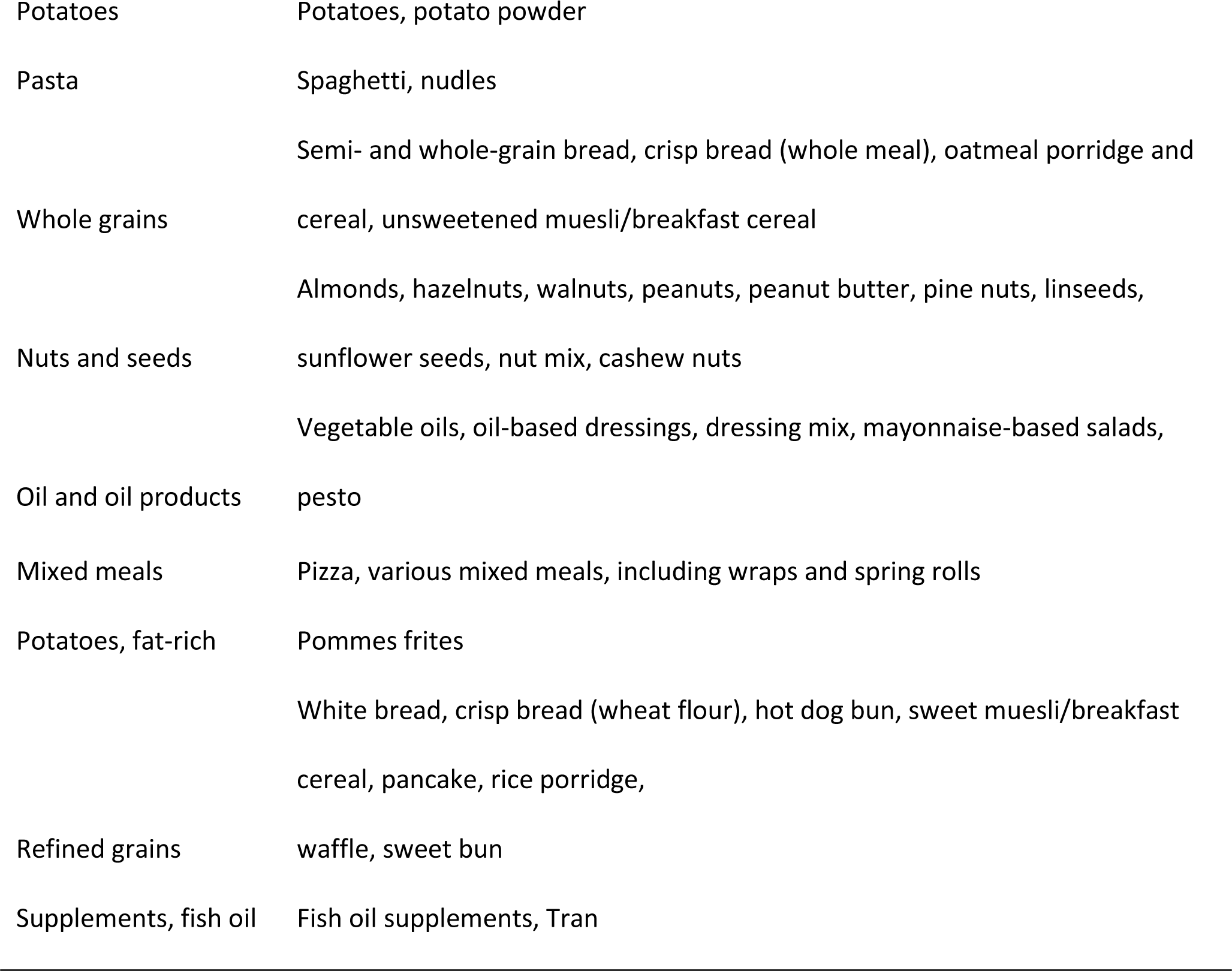
Groupings of food items used as input in the dietary pattern analysis.

See Supplemental Methods for an extended description of the data types.

### Statistical and bioinformatics analyses

Here we describe the statistical and bioinformatic analyses related to DPs, gene expression clusters, and statistical modeling. All analyses were performed in R version 3.6.2 (17). We refer to R packages and functions where appropriate, and using the following notation: `package::function`. Important deviations from default function setting are written in parentheses.

The flow of the analysis pipeline is outlined in **Supplementary Figure 1**. Women and men were analyzed separately, as preliminary analyses suggested a strong gender-related signal in both the DPs and gene expression dataset.

**Figure 1.**
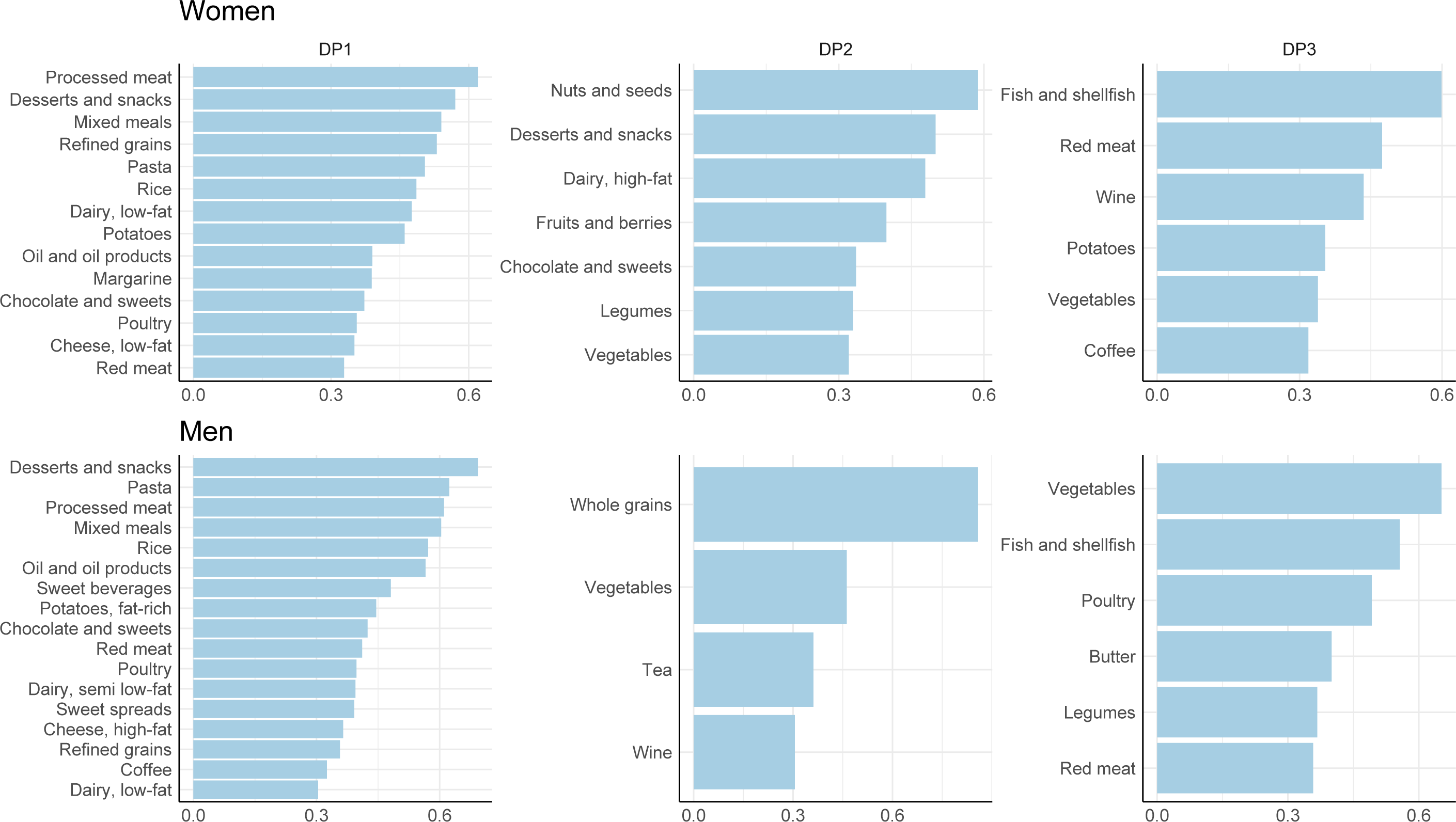
DPs for women and men. The figure shows factor loadings for all foods with a loading > 0.3. See Supplementary Table 4 for full Table of all foods (and their uniqueness). Abbreviations: DP, dietary pattern.

#### Dietary patterns

We used a combination of principal component analysis (PCA) and factor analysis to determine DPs. Factor analysis is a dimensionality reduction method similar to PCA, but it results in more interpretable features. However, because factor analysis is informed by the same covariance matrix as PCA, we used PCA-derived component variances (`stats::prcomp`) to determine a meaningful number of factors to retain; the results are presented in **Supplementary Figure 2**. For both genders, the eigenvalue-one criterion suggested around 12 principal components (PCs), but there was little change between components from component 3-5 and outwards; the scree test suggested around 3-5 components; the per component variance explained suggested that about 7-17 % of the variance could be explained until about three components, and then stabilized at 4-5 % at 4-6 components, with little change thereafter. We decided to extract three components using factor analysis (`stats::factanal`). The extracted DPs made sense from a dietary perspective (**Figure 1**), thus meeting the interpretability criterion.

**Figure 2.**
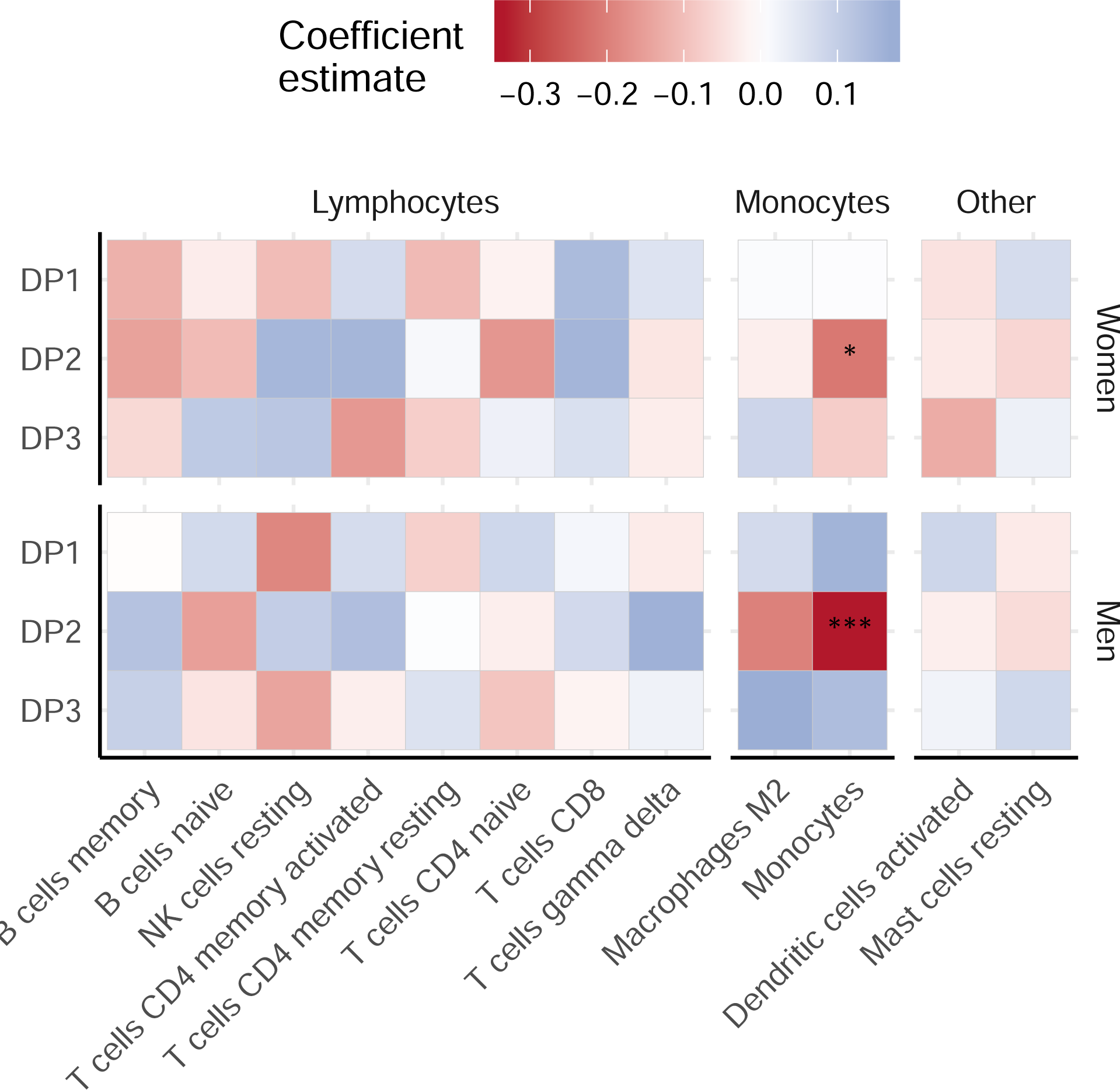
DPs associate with CIBERSORT-predicted cell types. The figure displays heatmaps of linear regression *β* coefficients between DP scores (as the exposure variable, shown in rows), and CIBERSORT-predicted cell types (as the outcome variable, in columns), for both women and men. Asterix indicate significance level: ***, P<0.001; **, P<0.01; *, P<0.05. See Supplementary Figure 1, Supplementary Figure 4 and Methods for a thorough explanation of the flow of analyses and adjustment levels. Abbreviations: DP, dietary pattern.

#### Gene expression features

Two main mechanisms are central in studies of diet-related associations with cardio-metabolic disease mechanisms in PBMCs: dietary effects on leukocyte subset distributions, and biological modulation independent of leukocyte subset distribution. As a result, we performed analyses to examine each of these aspects, as outlined in the upper right corner of Supplementary Figure 1.

##### Leukocyte subsets

We used CIBERSORT to perform *in silico* flow cytometry (13). This method uses support vector regression to conduct robust deconvolution of a heterogenous cell population, and returns predicted relative levels of various cell subsets. We used the raw, untransformed, whole-genome gene expression data matrix as input. Although the algorithm provides 22 leukocyte subsets, we filtered on the top most relevant cell types for the PBMC population, mainly monocyte and lymphocytes subsets, and thereby retained 12 cell subsets (**Supplementary Figure 3**). Note that although we had standard blood cell differential counts available, CIBERSORT resulted in a richer set PBMC cell subsets unique to the gene expression profile of each sample.

**Figure 3.**
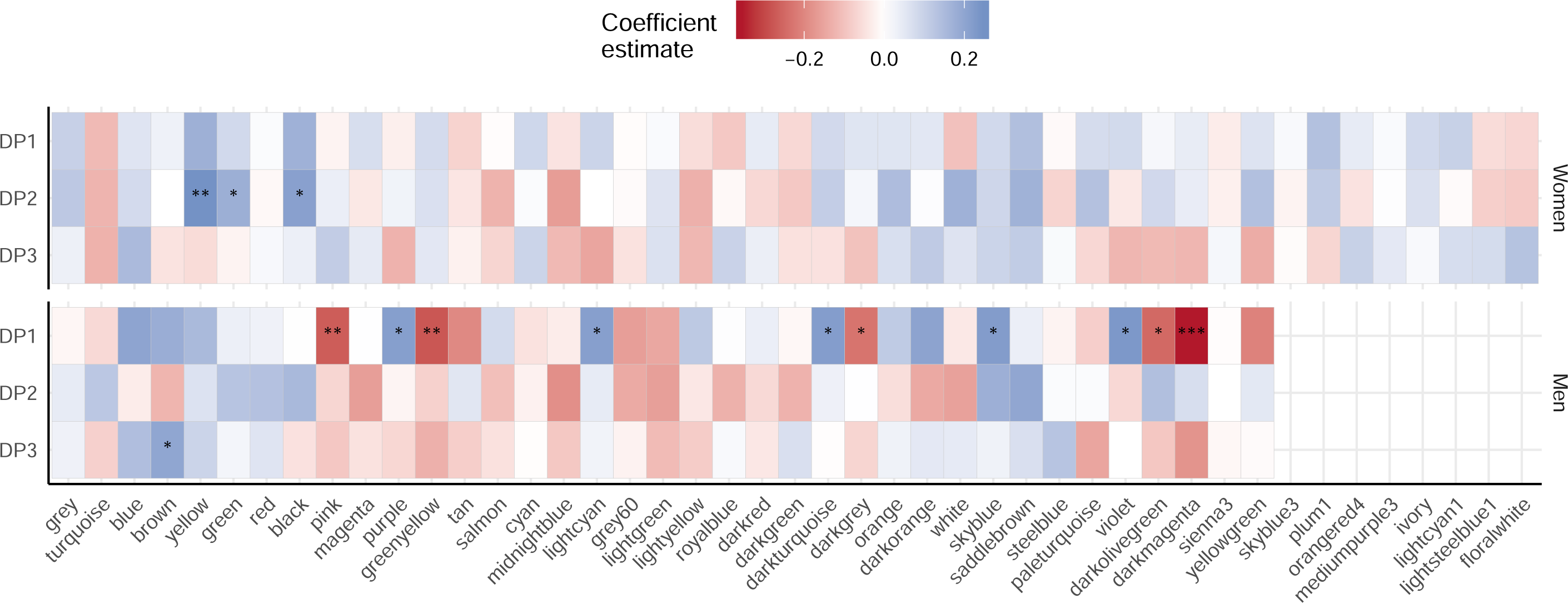
DPs associate with gene expression clusters. The figure displays heatmaps of linear regression *β* coefficients between DP scores (as the exposure variable, shown in rows), and gene expression cluster eigengenes (as the outcome variable, in columns), for both women and men. The clusters are sorted by size. Asterix indicate significance level: ***, P<0.001; **, P<0.01; *, P<0.05. See Supplementary Figure 1, Supplementary Figure 4 and Methods for a thorough explanation of the flow of analyses and adjustment levels. Abbreviations: DP, dietary pattern.

##### Gene expression clusters

We used WGCNA to identify highly correlated (“co-expressed”) clusters of genes (18). The `WGCNA` package (CRAN, Bioconductor) provides a well-established and popular framework to perform the WGCNA analysis (12). The details of the implementation can be found in (12); in Supplemental Methods we give a brief outline of key steps in the WGCNA-based gene expression cluster analysis pipeline.

To both avoid confounding by sex chromosomes and to aid compatibility with the dietary data, we performed the analysis separate for women and men. We examined the stability and validity of the resulting gene expression clusters between genders with module preservation statistics (19). Also, to avoid confounding by cell types, we adjusted for measured percentage monocytes and lymphocytes (standard differential counts), and extracted the residuals. Therefore, input for this analysis was the *residuals for the complete gene expression matrix* (p = 13967 variables)), after removing the main effect of monocytes and lymphocytes.

In order to highlight a few of the more important genes within interesting clusters, we performed a *driver gene* analysis. First, we calculated *cluster membership*, which is defined as the absolute correlation between gene expression and cluster eigengene, and can be interpreted as the degree to which each gene contributes to that cluster’s overall behavior, and contributes to its variation. Secondly, we calculated *DP significance*, which is the absolute correlation between gene expression and DP score. A positive correlation between cluster membership and DP significance indicates that those genes that drive the variation in the cluster eigengene are the same that drive the association with the specific DP (*driver genes*). Finally, to rank driver genes, driver gene estimates were calculated as the sum of the cluster membership and DP significance.

In order to describe relevant gene expression clusters biologically, we performed gene ontology (GO) enrichment using the GO Consortium database (20,21). Also, to link statistical findings with existing biological knowledge, we performed protein-protein interaction (PPI) network analyses using The Protein Interaction Network Analysis (PINA) 2.0 database (22).

#### Linear models

We associated DPs with two types of outcomes: CIBERSORT-predicted cell counts, and the eigengenes from the gene expression clusters using linear models (Supplementary Figure 1). **Supplementary Figure 4** shows the directed acyclic graphs (DAGs) used in model development. We used the open-access dagitty.net/dags web-resource to evaluate these relationships. Minimal sufficient adjustment sets for estimating the *total effect* of dietary pattern on gene expression were age and education (three levels: lower, middle, higher). For predicted cell counts, we additionally adjusted for adiposity (total fat mass, measured by bioelectrical impedance analysis) in sensitivity analyses. Also, in sensitivity analyses for the gene expression clusters, we estimated the *direct effect* (see Supplemental Methods).

**Figure 4.**
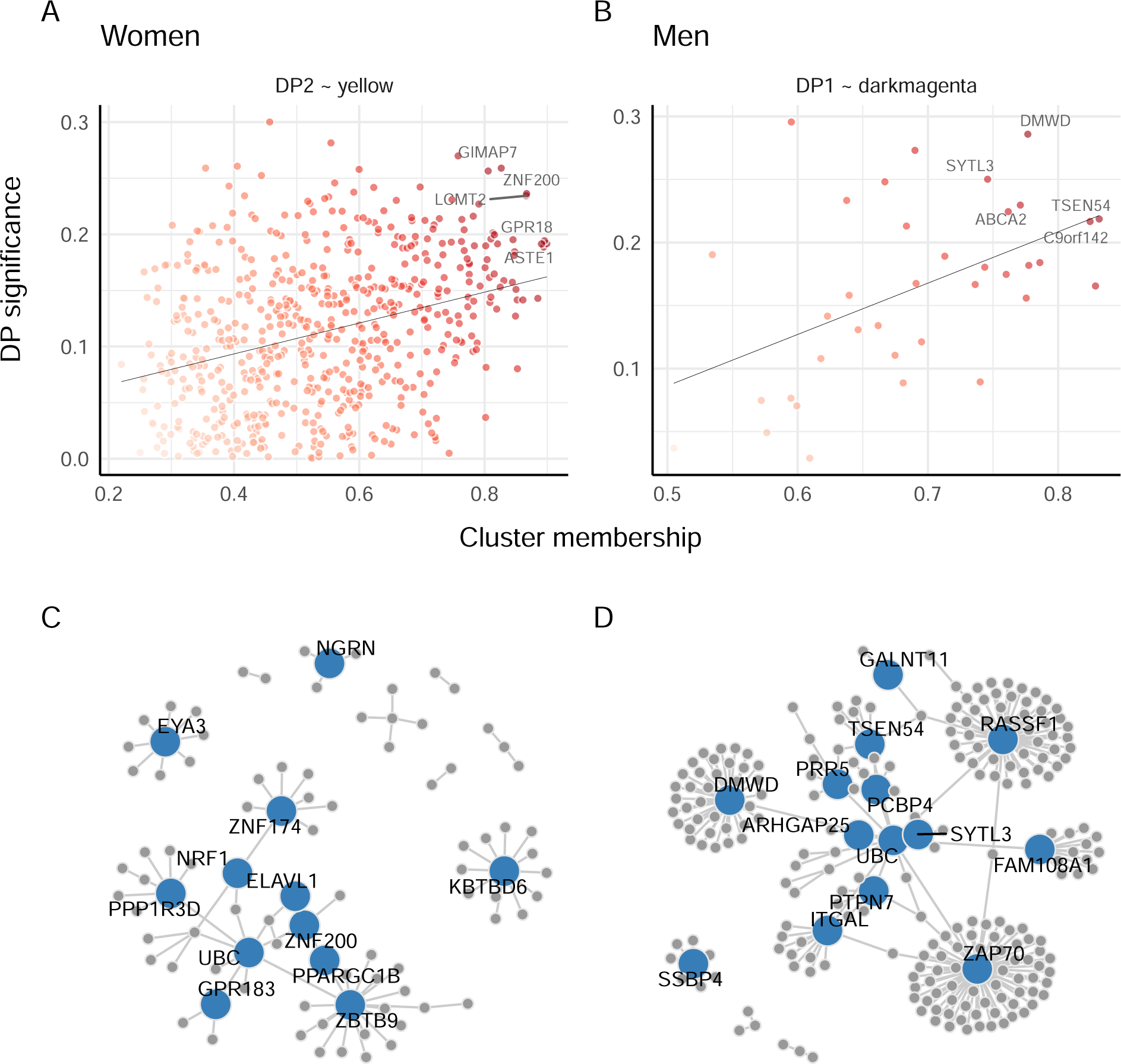
Identification of driver genes and hub proteins. A) and B) display the association between *cluster membership* and *DP significance* for the strongest DP and gene expression cluster associations, for each gender. Cluster membership is defined as the absolute correlation between gene expression and cluster eigengene, and can be interpreted as the degree to which each gene belongs in that certain cluster, and contributes to its variation. DP significance is the absolute correlation between gene expression and DP score. A positive correlation between cluster membership and DP significance indicates that those genes the drive the variation in the cluster eigengene are the same that associate with the specific DP (*driver genes*). Finally, to rank driver genes, driver gene estimates were calculated by the sum of the cluster membership and DP significance. The darker the color, the higher the driver gene estimate; the top five genes driving this association are annotated. Note strong positive correlations for both comparisons, as is also evident from the linear regression line. C) and D) show networks of protein-protein interactions (PPI) for the same DP and gene expression cluster associations as above. Each network was created by the top 20 driver genes identified by the driver gene plot. The figures display *hub proteins* that are of particular interest to the gene regulatory network. Abbreviations: DP, dietary pattern (see Supplementary Table 7 and Supplementary Table 8 for all abbreviations).

Note that for all models, technical covariates were considered in upstream batch correction (Supplementary Figure 1). Percentage of total leukocyte count of monocytes and lymphocytes (which make up the pool of PBMC subsets) were adjusted for in the gene expression pre-processing pipeline (as shown in Supplementary Figure 1), prior to WGCNA only. Finally, to aid interpretation of the results, we normalized (`base::scale`) both DP scores and cluster eigengenes to a standard normal distribution (mean = 0, sd = 1) before modeling.

#### Miscellaneous

The dietary intervention study was powered to detect a significant change in LDL-C (15); however, this does not apply to the present exploratory study. Because this study is exploratory, we did not evaluate associations by standard significance level cut-offs. Instead, we evaluated the strength and direction of associations, and their interrelations.

## Results

### Construction and description of study features

#### Dietary patterns

First, we constructed gender-specific DPs from self-reported FFQ data, yielding three DPs (Figure 1 and **Supplementary Table 4**). We considered the patterns to reflect typical *Western* (DP1), *Vegetarian* (DP2), and *Low carbohydrate* (DP3) diets. These three DPs explained 14.1, 8.0 and 6.6 %, and 16.6, 9.3 and 7.0 % of the variance, for women and men, respectively. Although there were some overlap, the *Vegetarian* and *Low carbohydrate* DPs were more unique to each gender compared to the *Western* DP. This was also supported by the DP loading for various foods (**Supplementary Figure 5**). For both genders, the *Western* DP associated with intake of meat and eggs, fastfood, snacks, dairy, and fiber-poor carbohydrate sources. The *Vegetarian* DP associated positively with a number of foods perceived as healthy, including plant foods, whole grains, nuts and seeds, and tea. Additionally, the association with animal products, fast food, dairy, and fiber-poor carbohydrate foods was low or negative. For women, the association with high-fat dairy and snacks was slightly positive. The *Low carbohydrate* DP was generally a mixture of the two former, reflected in positive associations for both plants and animal products. The association with fastfoods, snacks and carbohydrate-rich foods, however, was negative. Wine associated positively, whereas sweet beverages associated negatively with the *Low carbohydrate* DP for women and men, respectively.

In addition to the direct link with food intake, the DP scores correlated with both macronutrient intake (**Supplementary Figure 6**) and clinical variables (**Supplementary Figure 7**). The *Western* DP correlated with energy intake and negatively with fiber intake in both genders. The *Vegetarian* DP correlated positively with fiber and negatively with saturated fat intake in men. In women, the *Vegetarian* DP correlated weakly, but positively, with energy, healthy fats, fiber and sugar. The *Low carbohydrate* DP was negatively associated with carbohydrate and sugar intake in both genders, and with higher protein and fat intake in men.

For the clinical variables, the negative association between *Western* DP and age was most notable, which indicates that the younger part of the study sample adhere to a more unhealthy diet. Additionally, The *Vegetarian* DP associated negatively with multiple obesity-related markers, including immune cells and CRP. Again, the *Low carbohydrate* DP was a mixture of the two, with positive correlations for age and lipids.

#### Leukocyte subsets

We used the CIBERSORT algorithm to computationally estimate the distribution of 12 leukocyte subsets (13). As expected, predicted leukocyte cell proportions associated with multiple clinical variables, although most notably for the differential count measures and obesity-related measures (**Supplementary Figure 8**).

#### Gene expression clusters

Using the WGCNA algorithm, we detected 45 and 37 unique gene expression clusters for women and men, respectively, which by default were named different colors (12). Although there were large differences in cluster size (range = 67-307 and 85-438 genes for women and men, respectively), most clusters explained a large proportion of the variance of the genes they comprised (range = 32-39 and 33-40 % for women and men, respectively) (**Supplementary Figure 9A** and **B**, and **Supplementary Table 5**). For men, explained variance inversely associated with cluster size (**Supplementary Figure 9C**). In addition, genes in all clusters were generally distributed over all chromosomes, with certain exceptions, such as chromosome 1 and 19 (**Supplementary Figure 9D**). The gene expression clusters displayed some correlation within each gender, but they could largely be considered unique features (**Supplementary Figure 10**). Between genders, the module preservation was acceptable for most medium- and large-sized clusters, and poor for the smaller clusters (**Supplementary Figure 11, Supplementary Figure 12, Supplementary Figure 13**).

Numerous gene expression clusters correlated with clinical phenotypes (**Supplementary Figure 14**). Most prominent was the global associations with body composition- and lipid-related markers.

### Associations of derived gene expression features with dietary patterns

#### Dietary patterns and leukocyte subsets

Predicted percentage of monocytes associated negatively with the *Vegetarian* DP for both women (*β* = −0.21, *P* = 0.02) and men (*β* = −0.33, *P* = 0.0008) (**Figure 2** and **Supplementary Figure 15**), suggesting a link between this particular cell subset and diet. Interestingly, when adjusting for adiposity, this association was only attenuated for women (*β* = −0.15, *P* = 0.11 for women, and *β* = −0.33, *P* = 0.001 for men).

#### Dietary patterns and gene expression clusters

In general, relatively few associations were evident between DP scores and gene expression cluster eigengenes (**Figure 3**). For women, the positive association between the *Vegetarian* DP and the yellow cluster was strongest. The yellow cluster contained 600 genes involved in regulation of transcription (**Supplementary Figure 16**). For men, the *Western* DP associated with multiple clusters, of which the association with the darkmagenta cluster was strongest. This cluster contained 36 genes related to metabolic and inflammatory processes, including sterol/cholesterol transport (Supplementary Figure 16). Similarly, both the pink and greenyellow clusters associated negatively with the *Western* DP, although not as strongly as darkmagenta. The pink cluster consisted of 475 genes involved in regulation of viral processes, endosome/vacuolar transport, UDP-GlcNAc metabolism, and monocyte and lymphocyte stimulation. On the other hand, the greenyellow cluster consisted of 338 genes involved in regulation of protein synthesis and degradation, and acyl carnitine transport. The top 20 most enriched GO terms (for all three ontologies) for the top most significant cluster for each gender are listed in **Supplementary Table 6**.

### Identification of driver genes

Next, we examined the most relevant gene expression clusters more in detail, using a driver gene analysis to identify genes with both high DP significance and high cluster membership. Interestingly, DP significance and cluster membership associated strongly (**Figure 4A** and **B**, and **Supplementary Table 7**), which suggests that genes that associated with DPs were also among the most important parts of the clusters that associated with that DP.

The five top driver genes for the association between the *Vegetarian* DP and the yellow cluster in women were GIMAP7 (GTPase, IMAP family member 7), ZNF200 (zinc finger protein 200), LCMT2 (leucine carboxyl methyltransferase 2), GPR18 (G protein-coupled receptor 18), ASTE1 (asteroid homolog 1) (Figure 4A). Proteins from these genes regulate aspects of biosynthetic processes, including cell signaling, DNA transcription and repair, and protein synthesis (20,21). For these genes, the correlation coefficients with DP2 score were in the range 0.19 to 0.26 (*P* = - 0.003), and with the cluster eigengene in the range 0.83 to 0.90 (*P* < 0.001) (Supplementary Table 7). This means that women who consumed a *Vegetarian* DP tended to have *higher* expression of these genes in PBMCs.

The five top driver genes for the association between the *Western* DP and darkmagenta cluster in men were DMWD (DM1 locus, WD repeat containing), SYTL3 (synaptotagmin like 3), ABCA2 (ATP binding cassette subfamily A member 2), TSEN54 (tRNA splicing endonuclease subunit 54) and C9orf142/XLS (XRCC4-like small protein) (Figure 4B). Proteins from these genes are involved in lysosomal transport, cholesterol homeostasis, mRNA processing and DNA repair (20,21). The correlation coefficients with DP1 score were in the range −0.29 to −0.22 (*P* = 0.03 - 0.003), and with the cluster eigengene in the range 0.75 - 0.83 (*P* < 0.001) (Supplementary Table 7). This means that men who consumed a *Western* DP tended to have *lower* expression of these genes in PBMCs.

### Identification of hub proteins

Finally, to examine if these driver genes were part of physically interacting biological networks, we filtered them through the PINA database (22). For the strongest associations for each gender, we then created protein-protein interaction (PPI) networks (**Figure 4C** and **D**, and **Supplementary Table 8**). These proteins can be considered *hub proteins*; they likely exert a higher degree of control over the protein network, as more proteins physically interact with this hub in order to influence signaling pathways. For women and men, key hub proteins included PPARGC1B (PPARG coactivator 1 beta) and UBC (ubiquitin C), respectively.

## Discussion

In the present study of 235 Norwegian adults, we detected novel associations between DPs and gene expression features in PBMCs. Our results suggest that diet affects a number of specific cell types and pathways, of which the most pronounced are: predicted proportion of monocytes, regulation of transcription, and regulation of metabolic and inflammatory processes.

### We detected three DPs commonly consumed in Norway

Using data-driven analyses, we detected three DPs commonly consumed in Norway: *Western*-type, *Vegetarian*-type, and *Low carbohydrate*-type DP (Figure 1). These DPs were neither unexpected nor surprising: Norwegian adults follow trends, and this includes the vegetarian and low carbohydrate trends. In previous studies, similar names have been used to characterize the detected DPs. In a cohort of Norwegian postmenopausal women, Markussen and co-workers found four DPs, including the *Western* and *Vegetarian* DPs (23). In addition, they found a *High-protein* pattern that resembled our *Low carbohydrate* pattern. Their DPs, similar to ours, share characteristics and therefore also names, with DPs throughout Europe and the US. This emphasizes an important point: although the DPs retained in factor analyses are never exactly equal, as opposed to *a priori* methods, our three DPs share characteristics with many other DPs both in Norway and elsewhere (7,23–25).

The three DPs associated with food items, nutrient intake and clinical parameters to give a consistent picture of the DPs: in general, the *Low carbohydrate* DP appeared neutral compared to the *Western*-type and *Vegetarian*-type DPs, which associated with a number of unhealthy or healthy behaviors, respectively.

### Vegetarian DP associated with monocytes

The *Vegetarian* DP associated with CIBERSORT-predicted levels of monocytes (Figure 2), suggesting that gene expression related to monocyte differentiation and activity may be affected by diet. These results are corroborated by previous reports by others and us (26–29). Craddock and co-workers recently reviewed the evidence that vegetarian diets affect inflammatory and immune biomarkers, concluding that vegetarian diets associate with lower CRP, fibrinogen, and total leukocyte concentrations (26). Similarly, Eichelmann showed that plant-based diets cause reductions in obesity-related inflammatory biomarkers such as CRP, IL6 and sICAM (27). Indeed, our observed association between diet and monocyte level might be related to the degree of obesity in the population; however, we found only a slight attenuation of the association for women when adjusting for adiposity. In previous work, we have shown that both diet and risk factors may affect PBMC leukocyte distribution (28,29). We found that plasma omega 6 fatty acid level, as a marker of dietary intake of omega 6 fatty acids, associated with predicted leukocyte distribution (28). Vegetarian diets tend to have high content of vegetable oils, which may have affected our present results also. Similarly, we recently showed that children with familial hypercholesterolemia displayed an altered leukocyte distribution (29).

Most studies that examine the association between diet and immune cells use a modest number of established biomarkers, such as standard differential count or protein biomarkers. In the current analysis, however, we used approximately 14 000 mRNA transcripts from PBMCs, potentially making it a more sensitive test of associations with immune cell type distribution specifically, and inflammation in general (13). Additionally, our finding is important since it adds to the evidence that cell type distribution in cell mixtures can influence the association between diet and gene expression. This must be taken into account when interpreting PBMC gene expression results.

### DPs associated with few gene expression clusters

Few WGCNA-based gene expression clusters were associated with DPs, after correcting for variation in monocytes and lymphocytes number (Figure 3). This indicates that most of the co-variation between diet and gene expression in PBMCs relates to leukocyte cell type distribution. Nevertheless, in women, the *Vegetarian* DP associated most strongly with a cluster of genes involved in regulation of transcription, and in men, the *Western* DP associated most strongly with a cluster of genes related to metabolic and inflammatory processes, including sterol/cholesterol transport.

In previous reports, dietary intake of a healthy Nordic diet or omega-3 associated with expression of genes related to mitochondrial function, cell cycle, endoplasmic reticulum stress, apoptosis, and inflammatory processes (30–33). *Regulation of transcription* is another such unspecific term. Although highly unspecific, regulation of transcription may be a process related to age-related global or pathway-specific DNA methylation and gene expression (34–36). *Sterol/cholesterol transport*, on the other hand, is a highly specific biological process that is dramatically affected by diet and that affects disease risk (36,37). Plasma LDL-C is mainly determined by cellular sterol status and the functionality and activity of the LDL receptor; this in turn is a key determinant of disease risk (37). However, although cholesterol metabolism in liver and monocytes are tightly coupled and similarly regulated, our observed association likely results from molecular events occuring within the pool of PMBCs as they deal with cholesterol-related metabolic challenges. Nevertheless, PBMC expression of genes related to sterol/cholesterol transport could prove a robust marker of dietary variation (10).

Interestingly, the second and third most significant clusters that associated with the *Western* DP in men contained genes related to other metabolic processes, such as UDP-GlcNAc and acyl carnitine metabolism. While UDP-GlcNAc is involved in cellular glucose sensing, acyl carnitines are involved in fatty acid transport into the mitochondria (38,39). Indeed, in previous work, we found that plasma levels of acyl carnitines of specific chain lengths may be directly altered by changes in fatty acid quality of the diet (6). Taken together, these may be processes particularly sensitive to variation in dietary intake.

### We identified top driver genes and hub proteins

Finally, the WGCNA cluster analysis detected top driver genes that have been shown to physically interact in protein-protein interaction networks (Figure 4) (22). This is an important finding, as it provides further biological meaning to the statistical associations, and strengthens our belief that the top driver genes may be more than just spurious associations (40). The network analysis highlighted a few hub proteins that may act as central communicators within each cluster, such as UBC and PPARGC1B. The UBC protein is a key cell signaling molecule, especially related to ubiquitination, cytokine signaling, toll-like receptors, and nuclear factor kappa B (NFkB); in mouse models, knock-down of the ubiquitin system shows protection from diet-induced obesity (41). Furthermore, PPARGC1B is a transcriptional co-regulator involved in a number of biological processes, including thermogenesis, bone turnover and regulation of energy expenditure by fat and glucose oxidation. For example, Yin and co-workers recently showed that PPARGC1B affects PPAR alpha to protect against cardiomyophathy (42).

### Strengths and limitations

To the best of our knowledge, nobody has used CIBERSORT and WGCNA to study molecular associations with DPs. We believe these dimension reduction algorithms may be well suited to examine diet-related effects on PBMC cell type distribution using sensitive gene expression data. Although we have taken steps to minimize the probability of chance findings, we cannot rule this out completely. Our study sample is also relatively small, compared to for example Lin and co-workers (40). In addition to higher risk of false positive findings, low sample size also increases the risk of false *negative* findings, for example for the associations between DPs and WGCNA gene clusters. Furthermore, our study sample represents a highly selected part of the Norwegian population, limiting the generalizability of our results. In line with this limitation, our results should not be overinterpreted. Pathways related to metabolic regulation could potentially be biomarkers of dietary intake, and also potentially predict future risk (40,43).

However, more prediction research is needed before this can be realized.

## Conclusions

In conclusion, we detected novel associations between DPs and gene expression features in PBMCs. Our results suggest that DPs may affect monocytes proportions and regulation of biological processes, such as regulation of transcription and metabolic and inflammatory processes. Although the present findings are exploratory, our analysis pipeline serves as a useful framework for future studies on the association between diet and gene expression. More research is needed before our results can be translated into clinically meaningful biomarkers.

## Data Availability

The data will not be made publically available.

## Authorship

Conception and design: LFA, KBH, SMU. Data analysis: JJC, MT, KW. Data interpretation: JJC, MT, KBH, KW, LFA. Wrote paper: JJC, SMU, KW. In addition, the microarray hybridization and gene expression data pre-processing were performed at the Genomics Core Facility at NTNU, Trondheim, Norway.

## Abbreviations

DP: dietary pattern
PBMC: peripheral blood mononuclear cell
WGCNA: weighted gene co-expression network analysis
GO: gene ontology
PPI: protein-protein interaction

